# Role of MicroRNAs in Autosomal Dominant Polycystic Kidney Disease: Expression Profiles and Pathway Analysis

**DOI:** 10.1101/2024.10.01.24314548

**Authors:** Chandra Devi, Prashant Ranjan, Jitendra Kumar Mishra, Shivendra Singh, Parimal Das

## Abstract

This study investigates the role of specific microRNAs (miRNAs) in autosomal dominant polycystic kidney disease (ADPKD). Network analysis using Cytoscape 3.10.1 revealed 28 miRNAs that target two or more PKD related genes, while 14 miRNAs interacted with three or more genes. Pathway analysis of these 14 miRNAs, conducted via the miRPath V4.0 tool, utilized various resources such as KEGG, GO terms, Pfam, and MSigDB, highlighting their functional roles in ADPKD. Among these 14 miRNAs, four were prioritized for further analysis based on the higher number of target genes and existing evidence of their expression in blood. These miRNAs were analyzed for fold change expression levels using qRT-PCR on blood-derived RNA samples from ADPKD patients (n=19) and healthy controls (n=5). These four miRNAs—hsa-miR20a-5p, hsa-miR27b-3p, hsa-miR3613-3p, and hsa-miR181a-5p— showed significant upregulation with p-values of 0.0179, 0.0002, 0.0166, and 0.0402 respectively. The functional enrichment of miRNAs involved in PKD evidently suggests that these play diverse roles in regulating key processes such as cell cycle progression, mTOR signaling, autophagy, EMT, and cellular responses to hypoxia thus playing crucial role in the pathogenesis of ADPKD and could serve as potential biomarkers or therapeutic targets.

## INTRODUCTION

MicroRNAs (miRNAs) are usually about 22 nucleotides long small-noncoding RNAs that regulate gene expression by binding to the 3’ UTR of target genes. They play a crucial role in regulating wide range of cellular and biological processes including differentiation, apoptosis, proliferation, metastasis, embryogenesis, aging, and immunity (Goh *et al*., 2016; Tran and Montano, 2017; Shah and Shah, 2020). MiRNAs are produced through multiple processing events, including canonical processing from protein-coding transcriptional units and noncanonical processing from nonprotein-coding transcriptional units. While canonical intronic miRNAs rely on Drosha for processing and are co-transcriptionally processed with protein-coding transcripts, noncanonical intronic small RNAs, or mirtrons, bypass this step. Organized in clusters, miRNAs often target multiple mRNA transcripts within common cellular pathways, allowing for complex and adaptive regulatory control (Tran and Montano, 2017). While the mammalian genome encodes hundreds of miRNAs with organ-specific expression pattern, their expression in kidney tissue exhibits significant variation compared to other tissues and even within different segments of the kidney itself (Akkina and Becker, 2011). In recent years, research has revealed their significant involvement in numerous human diseases, encompassing cancer, diabetes, obesity, autoimmune diseases, cardiovascular diseases, and genetic disorders like polycystic kidney disease (PKD) (Ardekani and Naeini, 2010; Hajarnis, Lakhia and Patel, 2015). Studies have shown dysregulated expression of specific miRNAs in various stages of PKD development, suggesting their involvement in disease progression. For example, miRNAs including the miR17 ∼ 92 cluster, miRNA-21, miR-15a, miR-200 family, and miR-199a, have been identified as key regulators of cyst growth, fibrosis, and epithelial-to-mesenchymal transition (EMT) and disease pathogenesis in autosomal dominant PKD (ADPKD) (Li and Sun, 2020; Lai *et al*., 2024). The altered expression of miRNAs such as miR-192, miR-204, and miR-509-3p has been associated with renal cyst formation and disease severity in both animal models and human patients. Epigenetic regulation presents another potential biological mechanism that may elucidate the heterogeneity in ADPKD. Epigenetic changes, encompassing histone modifications like acetylation, methylation, and phosphorylation, have been implicated in the pathogenesis of ADPKD, with increased TGF-β levels observed to induce DNA methylation in ADPKD tissue, potentially contributing to kidney fibrosis (Norman, 2011; Fontecha-Barriuso *et al*., 2018). MiRNAs play a significant role in ADPKD by directly binding to target genes and influencing cellular processes such as proliferation, differentiation, and apoptosis. While targeting miRNAs that bind to *PKD1* or *PKD2* holds promise for regulating clinical manifestations, it is recognized that miRNAs alone may not suffice for treating ADPKD and additional therapeutic strategies are required. With the identification of various miRNAs involved in cystogenesis through microarray data, research on miRNAs in ADPKD is anticipated to become increasingly active in the future. A promising approach involves the development of a new class of therapeutics known as antimiRs and miRNA mimics, designed to modulate miRNA activity. These drugs are undergoing pre-clinical and clinical evaluations (Li and Sun, 2020), raising the prospect of miRNA-based interventions for common human diseases. Through a miRNA enrichment and pathway analysis, this study aims to analyze the involvement of specific miRNAs in ADPKD.

## METHODOLOGY

### Study participants and ethical approval

The study was approved by the Ethics Committee of the Institute of Science, Banaras Hindu University, India. A total of 19 ADPKD patients were recruited from Department of Nephrology, Sir Sunderlal Hospital, Banaras Hindu University, with informed consent obtained from all individuals. Peripheral whole blood samples from 5 healthy controls were voluntarily provided by research laboratory members after informed consent. To maintain confidentiality and anonymity, each sample was assigned a unique study code.

### MiRNA shortlisting

Literature search was done on PubMed using the search query ‘miRNA and Polycystic Kidney Disease.’ A list of miRNAs was extracted mentioned within selected articles (Table 1 in Supplementary file 1). Any duplicate miRNAs were removed resulting in a concise set of miRNAs associated with PKD.

**Table 1:**
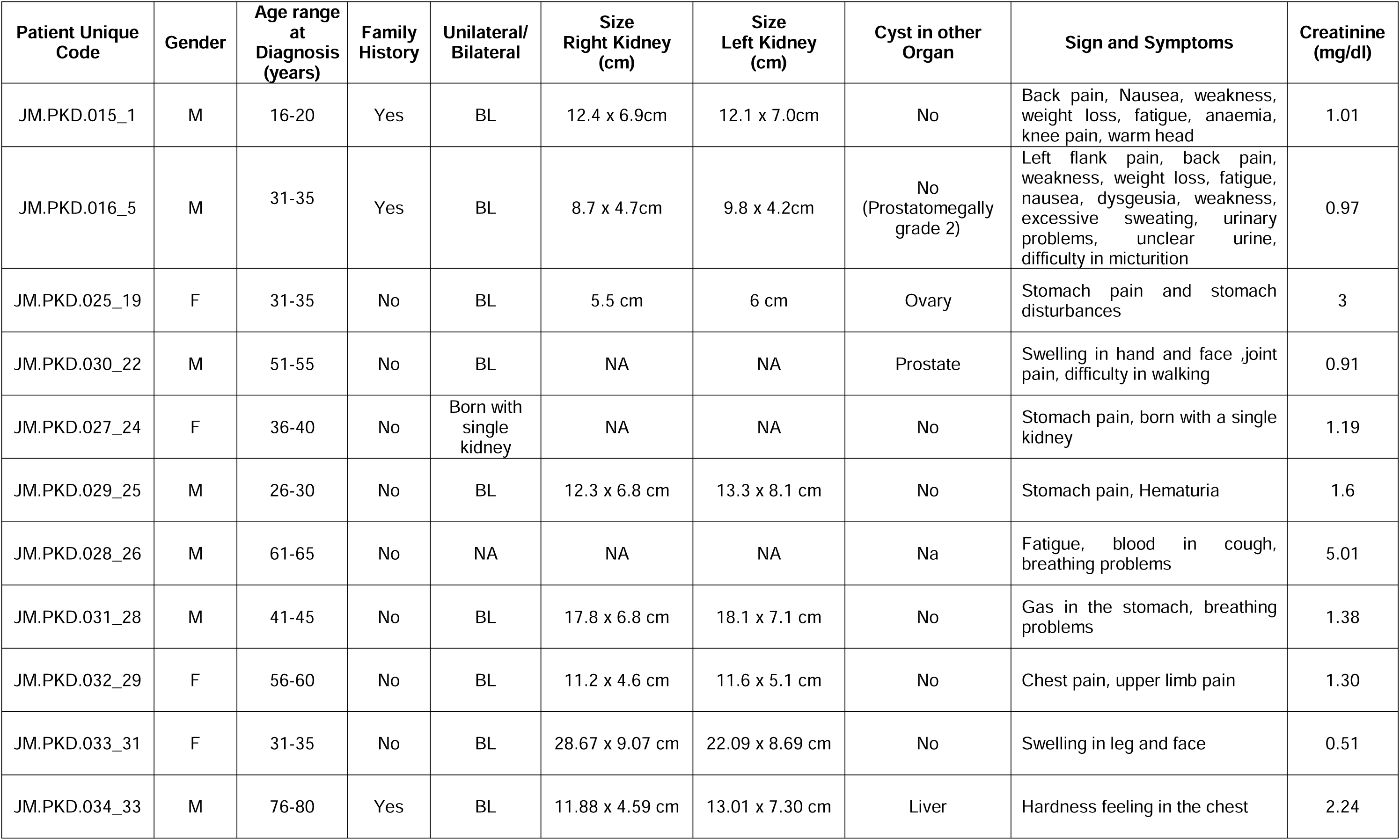
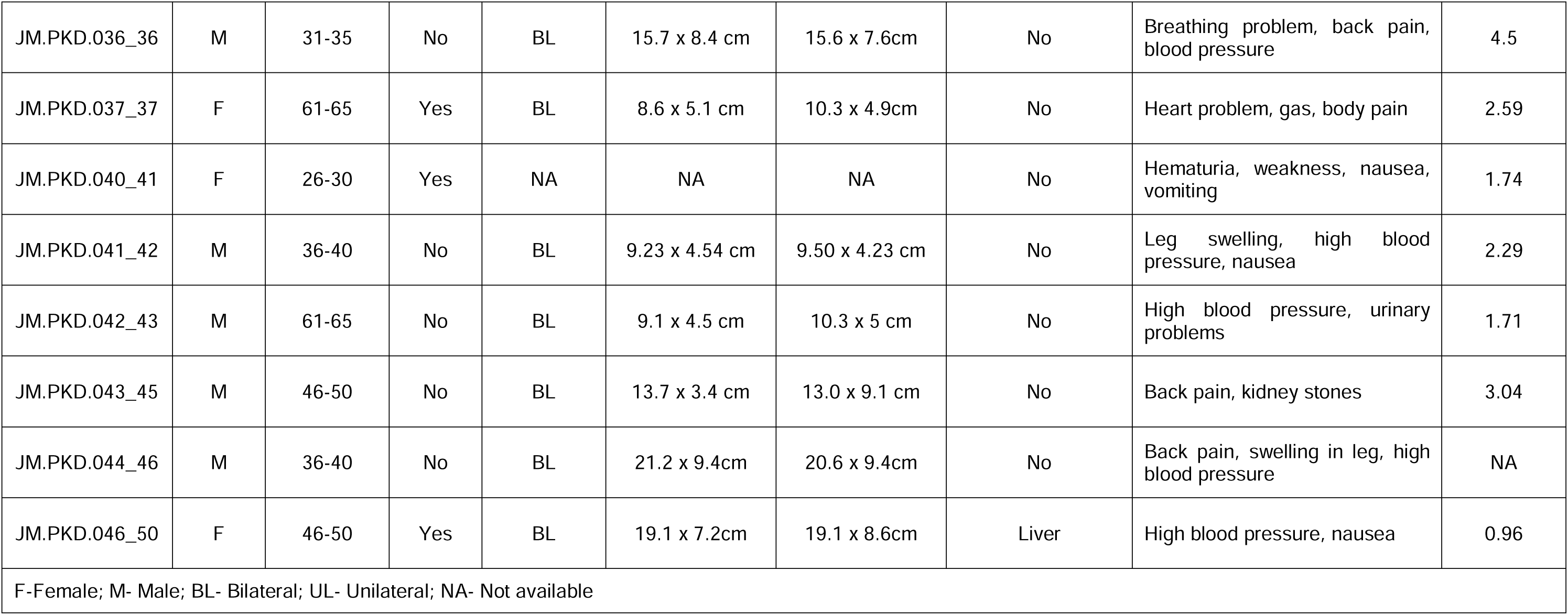
Clinical Characteristics of the Study Participants with Autosomal Dominant Polycystic Kidney Disease.

### Retrieval of Target Genes

To investigate the regulatory network of these miRNAs, miRDB database was accessed, a database for miRNA target gene information (Chen and Wang, 2020). A threshold of target values equal to or exceeding 90 was considered for selecting the target genes. This stringent criterion allowed curating a list of miRNAs that play a role in the context of PKD.

### Selection of PKD-Associated Genes

Two databases, GeneCard and OMIM, were searched using the term ‘Polycystic kidney disease,’ and a concise list of 108 genes that were common to both databases was compiled.

### Mapping miRNA-Target Gene Interactions

With the lists of miRNAs with their target genes and PKD-associated genes, we proceeded to identify interactions between miRNAs and their respective target genes within the context of PKD. The target genes of each miRNA were searched within the list of 108 PKD-associated genes and common genes between miRNA target genes and PKD-associated genes were identified. Thus a final list of 64 miRNAs and their corresponding common target genes was compiled serving as the basis for the subsequent analysis.

### miRNA-Target Gene Network and Functional Enrichment

Cytoscape (version 3.10.1) was used to visualize and explore the relationships between the identified miRNAs and their target genes and for the functional enrichment of the associated genes (Kohl, Wiese and Warscheid, 2011).

### Pathway Analysis for miRNAs

Pathway analysis was carried out using the miRPath V4.0 tool (Vlachos *et al*., 2012), an online resource available through the Diana Tool to understand the biological implications of the 14 miRNAs that exhibited interactions with three or more genes. Various pathway resources were used, including KEGG, GO terms (Cellular Component, Biological Process, and Molecular Function), Pfam, and MSigDB, to gain insights into the functional roles of these miRNAs.

### RNA isolation

RNA was isolated from whole blood using TRIzol reagent (Invitrogen) according to the manufacturer’s instructions. RNA quality and quantity were evaluated using spectrophotometric analysis (Nanodrop 2000, Thermo Scientific Inc.). The RevertAid First Strand cDNA Synthesis Kit (Catalog #K1622, Thermo Scientific, USA) was utilized for reverse transcription of miRNAs from the total RNA using specific stem-loop primers.

### Stem-loop RT-qPCR Analysis

For the functional analysis of miRNAs, Stem-loop RT-qPCR primers were designed for the selected four miRNAs using the sRNAPrimerDB online tool (Supplementary Table 2). These miRNAs were selected based on the higher number of target genes and existing evidence of their expression in blood. The miRNA nucleotide sequences were obtained from the miRBase database. Reverse transcription (total volume = 10 μL) was performed on 200 ng of RNA with miRNA-specific stem-loop primers. Quantitative PCR (qRT-PCR) was conducted on the Applied Biosystems QuantStudio 6 Flex Real-Time PCR System (Thermo Scientific, USA). U6 small nuclear RNA was used as the endogenous control for normalizing target miRNA expression. The reaction mix included 1 μL of cDNA (pre-diluted 1:5), 6.25 μL of Maxima SYBR Green/ROX qPCR Master mix (2X, Thermo Scientific, USA), 1 μL of miRNA-specific forward primer, 1 μL of reverse primer (both at 10 μM), and nuclease-free water for a final volume of 12.5 μL. The amplification conditions consisted of an initial denaturation at 95°C for 10 minutes, followed by 40 cycles of 95°C for 15 seconds and 60°C for 1 minute. Quantification was achieved using comparative CT (ΔCT) analysis, with fold change determined accordingly.

### Statistical analysis

Statistical data are expressed as mean ± SEM. Analyses were conducted using GraphPad Prism 10 software, and statistical significance was determined using a two-tailed unpaired Student’s t-test. The p-value of less than 0.05 was considered statistically significant.

## RESULTS

### MiRNA-Target Gene Interactions

In our network analysis of 64 miRNAs, we explored the interactions between miRNAs and their target genes. This analysis revealed 40 unique target genes, each interacting with different miRNAs, forming a regulatory network associated with PKD (Figure 1). In this network, 28 miRNAs targeted to two or more genes and 14 miRNAs have interactions with three or more target genes. The 14 genes and their respective target gene are listed in Table 2. Among these 14 miRNAs, four miRNAs viz., hsa-miR-20a-5p, hsa-miR-27b-3p, hsa-miR-3613-3p, and hsa-miR-181a-5p, were selected for expression analysis in blood samples based on the higher number of target genes and existing evidence of their expression in blood.

**Figure 1:**
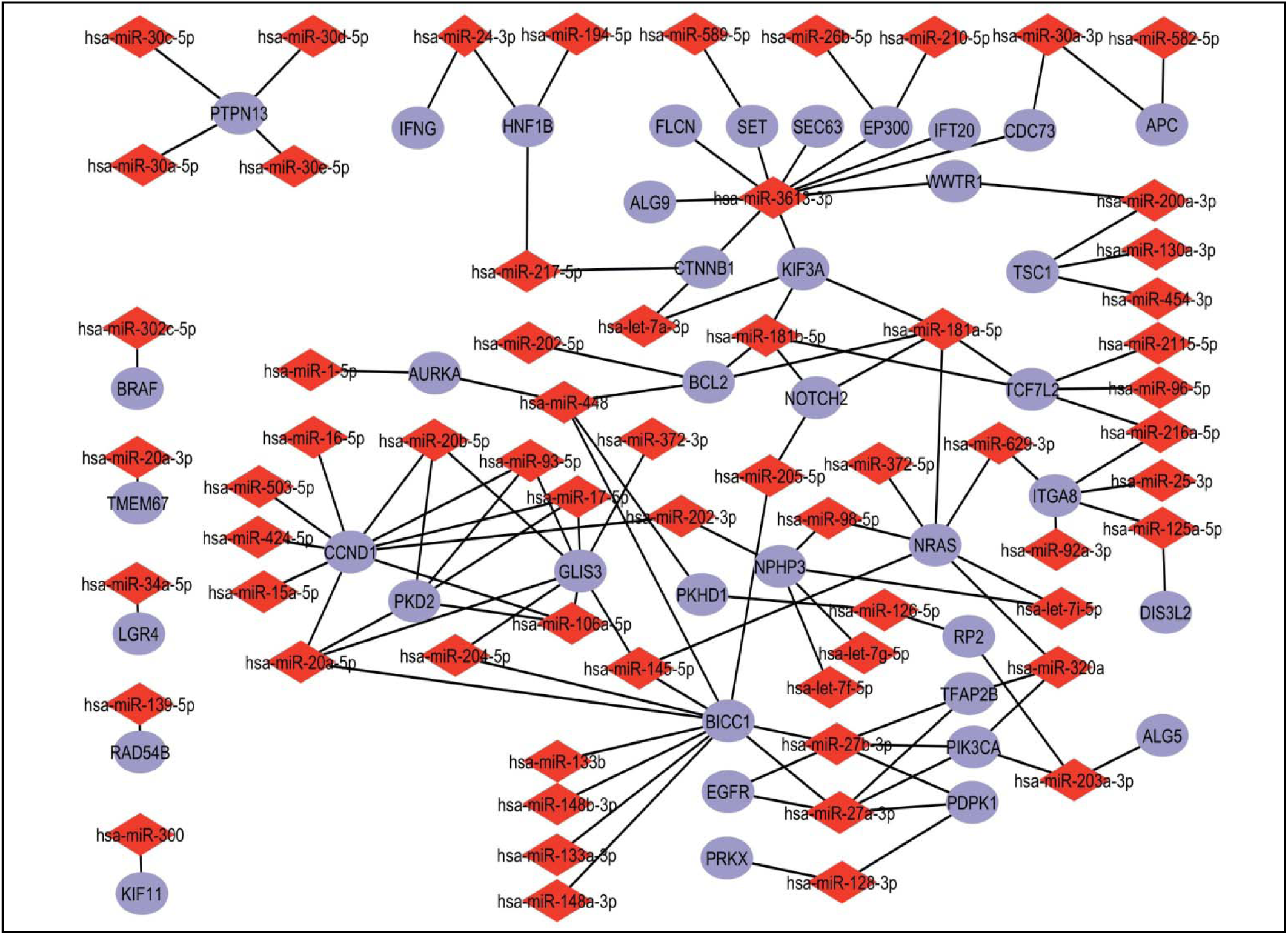
Network of miRNA-Target Gene Interactions in ADPKD: The network analysis was performed using Cytoscape 3.10.1 to map the interactions between 64 miRNAs and their target genes. The visualization illustrates the regulatory relationships. Twenty eight miRNAs target two or more genes and 14 miRNAs interact with three or more genes. Nodes represent miRNAs (red) and target genes (blue), while edges depict their interactions.

**Table 2:**
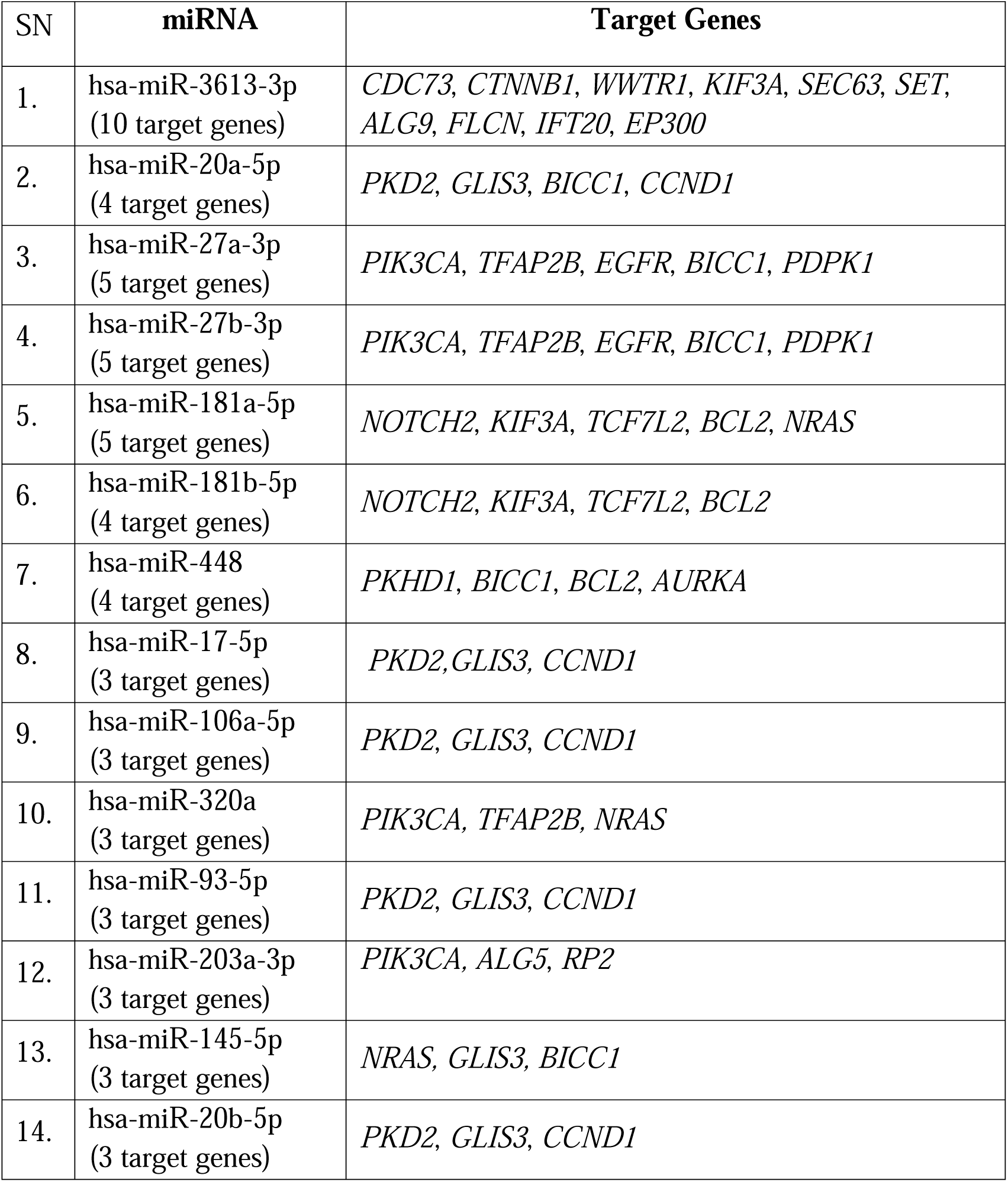
List of 14 miRNAs that target three or more genes.

| SN | miRNA | Target Genes |
| --- | --- | --- |
| 1. | hsa-miR-3613-3p<br>(10 target genes) | *CDC73, CTNNB1, WWTR1, KIF3A, SEC63, SET, ALG9, FLCN, IFT20, EP300* |
| 2. | hsa-miR-20a-5p<br>(4 target genes) | *PKD2, GLIS3, BICC1, CCND1* |
| 3. | hsa-miR-27a-3p<br>(5 target genes) | *PIK3CA, TFAP2B, EGFR, BICC1, PDPK1* |
| 4. | hsa-miR-27b-3p<br>(5 target genes) | *PIK3CA, TFAP2B, EGFR, BICC1, PDPK1* |
| 5. | hsa-miR-181a-5p<br>(5 target genes) | *NOTCH2, KIF3A, TCF7L2, BCL2, NRAS* |
| 6. | hsa-miR-181b-5p<br>(4 target genes) | *NOTCH2, KIF3A, TCF7L2, BCL2* |
| 7. | hsa-miR-448<br>(4 target genes) | *PKHD1, BICC1, BCL2, AURKA* |
| 8. | hsa-miR-17-5p<br>(3 target genes) | *PKD2, GLIS3, CCND1* |
| 9. | hsa-miR-106a-5p<br>(3 target genes) | *PKD2, GLIS3, CCND1* |
| 10. | hsa-miR-320a<br>(3 target genes) | *PIK3CA, TFAP2B, NRAS* |
| 11. | hsa-miR-93-5p<br>(3 target genes) | *PKD2, GLIS3, CCND1* |
| 12. | hsa-miR-203a-3p<br>(3 target genes) | *PIK3CA, ALG5, RP2* |
| 13. | hsa-miR-145-5p<br>(3 target genes) | *NRAS, GLIS3, BICC1* |
| 14. | hsa-miR-20b-5p<br>(3 target genes) | *PKD2, GLIS3, CCND1* |

Genes that are targeted by multiple miRNAs are *CCND1, NRAS, GLIS3, BICC1, KIF3A, TCF7L2*. These genes, targeted by multiple miRNAs, may play critical roles in the regulatory pathways that contribute to ADPKD pathogenesis. The convergence of miRNAs on these genes highlights their importance as molecular hubs, possibly involved in key processes such as cell cycle regulation (e.g., *CCND1*), signal transduction (e.g., *NRAS*), and ciliary function (e.g., *KIF3A*). Multiple miRNAs target common genes within these pathways further strengthens the notion that miRNA-mediated dysregulation plays a crucial role in ADPKD. Therapeutically, these genes and their interacting miRNAs present potential targets for modulating disease progression in ADPKD.

### MiRNAs expression analysis from blood

Fold change expression analysis of four miRNAs using Stem-loop RT-qPCR was performed on RNA extracted from whole blood samples of ADPKD patients (T) and healthy controls (C). The clinical characteristics of the ADPKD participants are presented in Table 1. The results revealed significantly higher expression levels of these miRNAs in ADPKD patients compared to healthy controls. The p-values for hsa-miR20a-5p, hsa-miR27b-3p, hsa-miR3613-3p, and has-miR181a-5p were 0.0179 (C = 5, T = 19), 0.0002 (C = 5, T = 18), 0.0166 (C = 5, T = 15), and 0.0402 (C = 5, T = 15) respectively (Figure 2).

**Figure 2:**
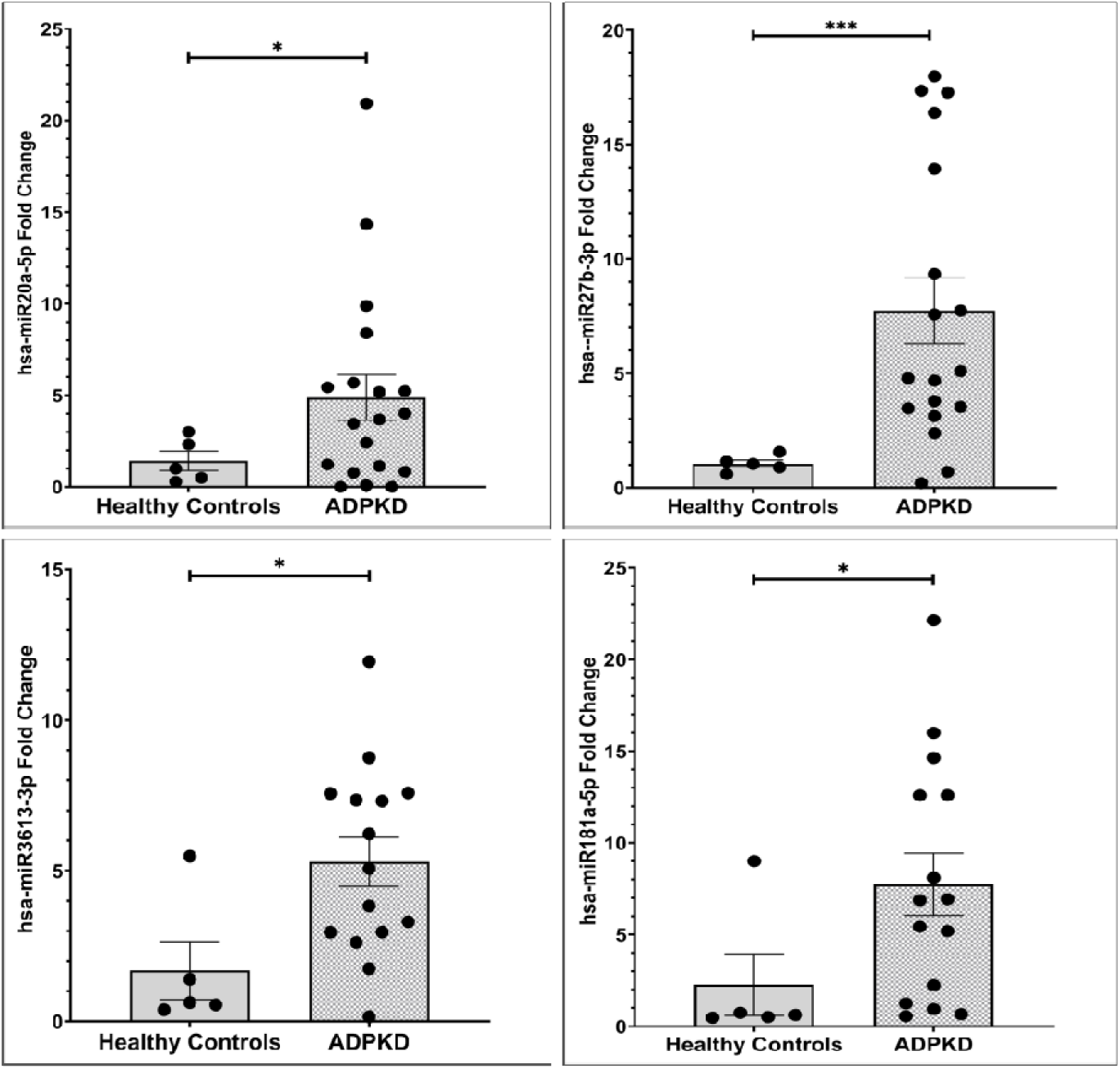
Fold change expression analysis using RT-qPCR from blood-derived total RNA. The fold change in expression levels of hsa-miR20a-5p, hsa-miR27b-3p, hsa-miR3613-3p, and has-miR181a-5p represent the higher expression of these microRNAs in ADPKD patients compared to healthy controls. The p-values for hsa-miR20a-5p, hsa-miR27b-3p, hsa-miR3613-3p, and has-miR181a-5p were 0.0179 (C = 5, T = 19), 0.0002 (C = 5, T = 18), 0.0166 (C = 5, T = 15), and 0.0402 (C = 5, T = 15) respectively. Data are depicted as mean ± SEM. *p < 0.05, ***p < 0.001, compared to healthy control group determined by two-tailed unpaired Student’s t-test.

### Pathway Analysis for miRNAs

The pathway analysis of the 14 miRNAs utilizing KEGG, Gene Ontology (GO) terms (cellular component, biological process, molecular function), Pfam, and MSigDB Hallmark Pathways identified multiple significant pathways and processes potentially implicated in ADPKD. KEGG analysis highlighted several relevant pathways such as ubiquitin-mediated proteolysis, focal adhesion, ErbB signaling, Hippo signaling, and mTOR signaling, which are known to be involved in cell proliferation, apoptosis, and tissue remodeling—all critical processes in cyst formation and expansion in ADPKD. Pathways such as cell cycle regulation, protein ubiquitination, and autophagy were particularly enriched, aligning with known mechanisms of ADPKD pathogenesis, where dysregulated cell cycle and impaired protein degradation contribute to abnormal epithelial cell growth and cystogenesis (Figure 3).

**Figure 3:**
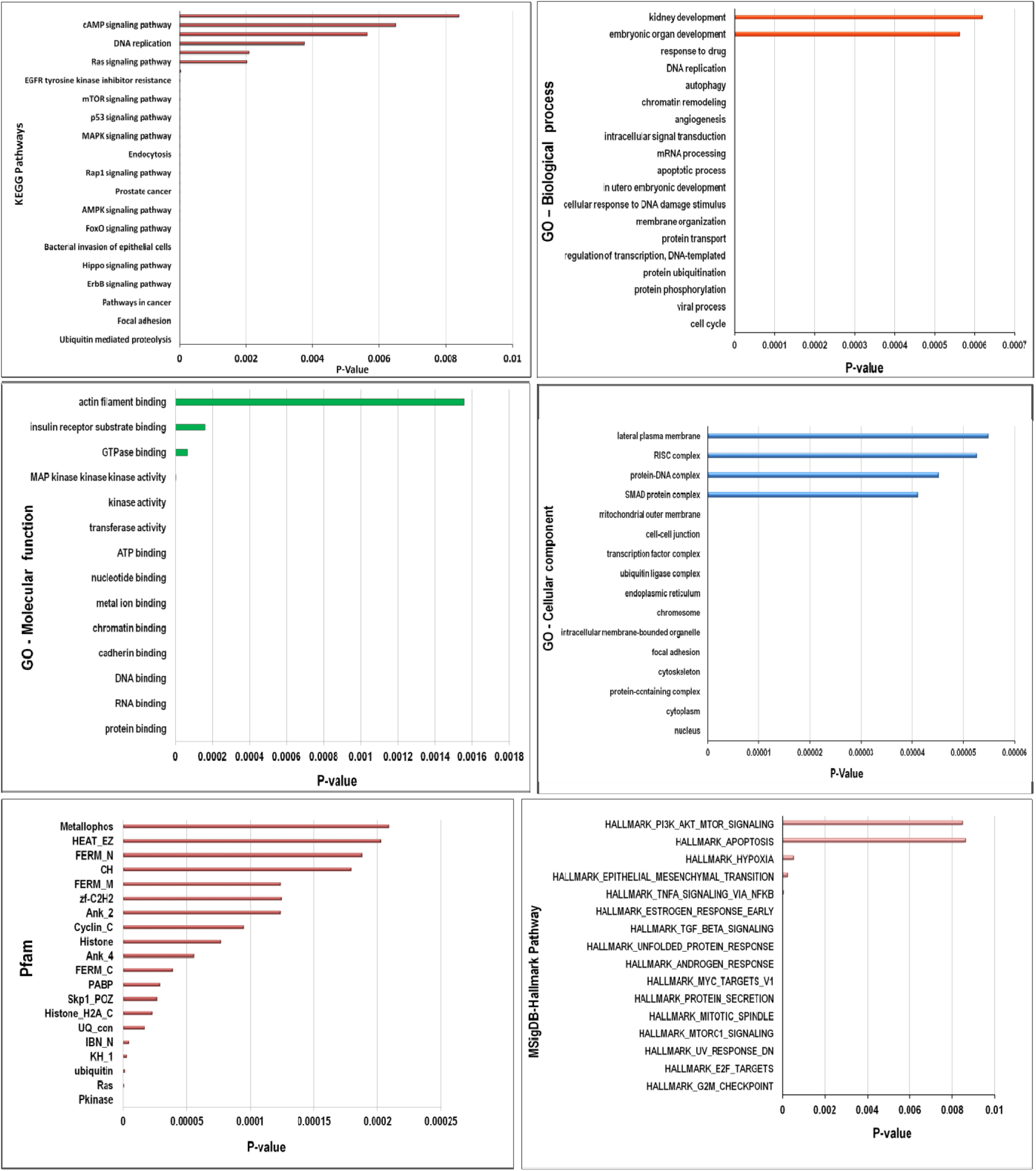
Pathway Analysis of miRNAs with Multiple Gene Interactions. Pathway analysis was conducted on 14 miRNAs that exhibited interactions with three or more genes. The analysis incorporated various pathway resources, including KEGG, GO terms (encompassing Cellular Component, Biological Process, and Molecular Function), Pfam, and MSigDB suggesting possible functional roles and biological significance of these miRNAs in ADPKD.

GO cellular component analysis revealed enrichment in pathways associated with the nucleus, cytoplasm, protein-containing complexes, and cytoskeleton, indicating miRNA regulation of cellular structures essential for maintaining epithelial cell integrity (Figure 3).

Biological processes such as cell cycle regulation, protein phosphorylation, ubiquitination, DNA damage response, intracellular signaling, and apoptosis were highly represented, with significant enrichment in pathways like the G2M checkpoint, E2F targets, and mTORC1 signaling, all of which are known to contribute to epithelial cell proliferation and cyst expansion (Figure 3).

The molecular function analysis pointed to protein binding, RNA binding, DNA binding, and kinase activity, suggesting miRNA-mediated regulation of signaling pathways (Figure 3).

The Pfam analysis identified domains like Pkinase, Ras, ubiquitin, and histone-related families, further supporting the involvement of miRNAs in protein kinase activity, ubiquitination processes, and chromatin remodeling in the context of ADPKD (Figure 3).

The MSigDB Hallmark pathways such as G2M checkpoint, E2F targets, unfolded protein response, TGF-beta signaling, mTORC1 signaling, and EMT were also enriched, indicating a role for miRNAs in regulating cyst growth, fibrosis, and cellular stress responses in ADPKD (Figure 3).

### Functional enrichment analysis

From the **functional enrichment analysis** of miRNA target genes, we identified 33 genes associated with renal cyst formation. These genes are: *TSC2*, *PKD2*, *B9D2*, *NOTCH2*, *PKD1*, *INVS*, *NEK8*, *FLCN*, *BUB1B*, *LRP5*, *ANKS6*, *TSC1*, *ALG8*, *BUB1*, *STK11*, *DZIP1L*, *NPHP3*, *GANAB*, *OFD1*, *CDC73*, *SEC63*, *PKHD1*, *GLIS3*, *TMEM67*, *CDKN1C*, *DNAJB11*, *NEK1*, *AKT1*, *ZNF423*, *PRKCSH*, *HNF1B*, *ALG9*, *MUC1* (Figure 4A and Supplementary file 2). Further the top 20 hub genes in the network are *NOTCH2, CCND1, IFNG, TP53, TSC2, PIK3CA, EGFR, SRC, CTNNB1, STK11, NRAS, CASP3, AKT1, BCL2, TNF, MUC1, TLR2, STAT6, EP300,* and *BRAF* (Figure 4B). These genes are integral to understanding the molecular mechanisms underlying renal cyst development.

**Figure 4:**
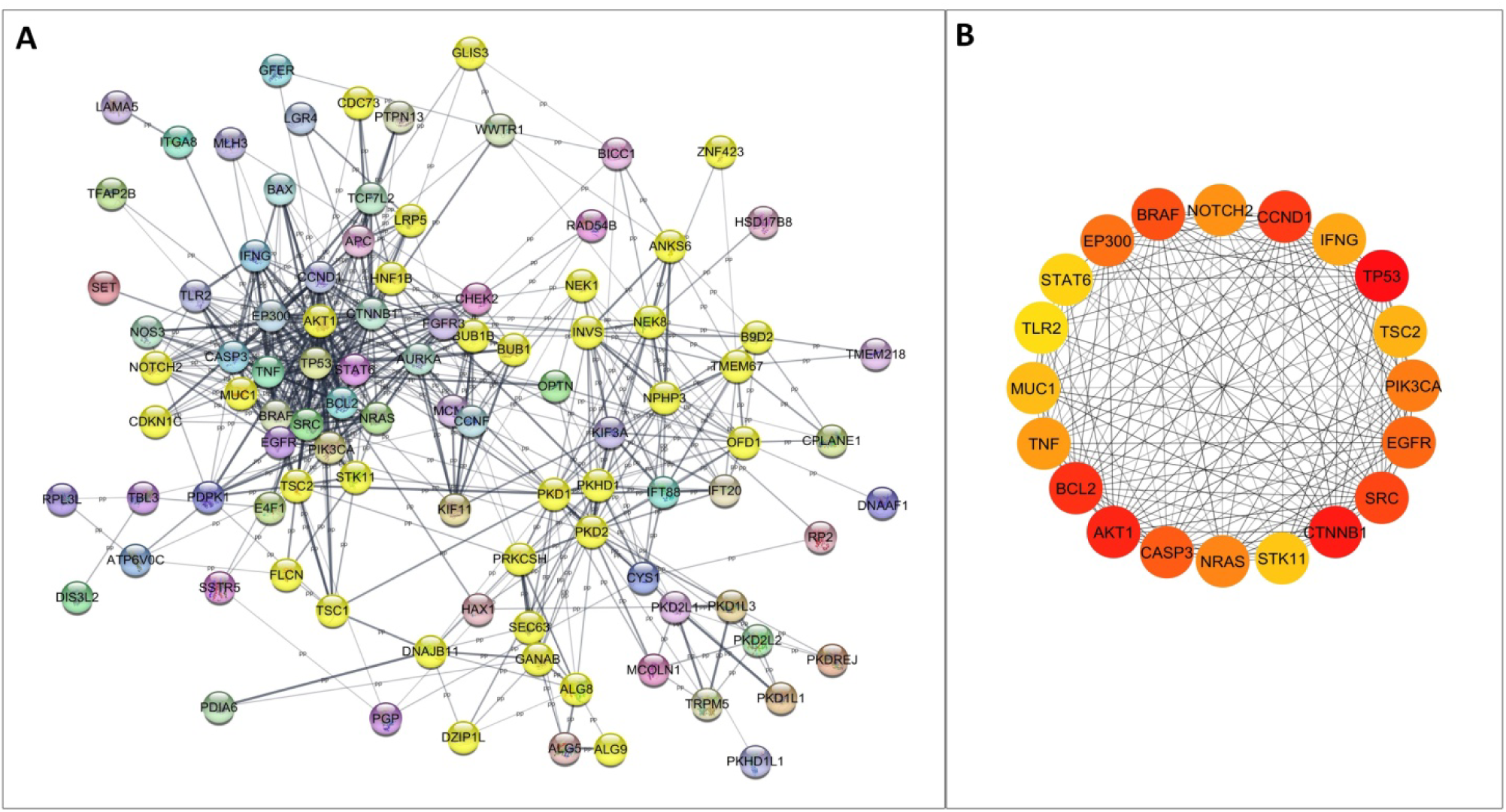
Functional Enrichment Analysis of miRNA-Target Genes. This network visualization, generated using Cytoscape 3.10.1, represents **A)** the target genes regulated by 64 miRNAs and common list of PKD-associated genes from GeneCard and OMIM. The functional enrichment analysis pinpointed 33 genes specifically linked to renal cyst development, which are also highlighted in yellow. These are: *TSC2*, *PKD2*, *B9D2*, *NOTCH2*, *PKD1*, *INVS*, *NEK8*, *FLCN*, *BUB1B*, *LRP5*, *ANKS6*, *TSC1*, *ALG8*, *BUB1*, *STK11*, *DZIP1L*, *NPHP3*, *GANAB*, *OFD1*, *CDC73*, *SEC63*, *PKHD1*, *GLIS3*, *TMEM67*, *CDKN1C*, *DNAJB11*, *NEK1*, *AKT1*, *ZNF423*, *PRKCSH*, *HNF1B*, *ALG9*, *MUC1.* **B)** The top 20 hub genes identified within the miRNA-target gene interaction network. These hub genes demonstrate a central role in context of ADPKD.

## DISCUSSION

In this study, we investigated miRNAs implicated in PKD by identifying their target genes. The miRNAs with PKD-related target genes were identified, followed by enrichment and pathway analysis to explore their biological roles. The expression levels of four key miRNAs in ADPKD patients were assessed using qRT-PCR, revealing significant dysregulation associated with disease pathogenesis. ADPKD, a common genetic renal disorder, is characterized by the gradual formation of renal cysts, leading to kidney enlargement and eventual failure. Despite extensive research on the primary genetic mutations in *PKD1* and *PKD2*, the molecular mechanisms driving cystogenesis remain unclear. Emerging evidence points to miRNAs as crucial post-transcriptional regulators of gene expression and signaling pathways involved in cell proliferation, apoptosis, and tissue remodeling—processes central to ADPKD.

Expression analysis of four of the key miRNAs—hsa-miR-20a-5p, hsa-miR-27b-3p, hsa-miR-3613-3p, and hsa-miR-181a-5p—revealed significantly elevated levels in the blood samples of ADPKD patients compared to healthy controls. The miRNA-target gene network analysis revealed that these miRNAs regulate multiple essential cellular processes linked to ADPKD pathogenesis. For example, miR-20a-5p targets *PKD2*, *GLIS3*, *BICC1*, and *CCND1*, which are involved in pathways associated with cell cycle regulation and cyst development. The *PKD2, GLIS3,* and *CCND1* are also the target of hsa-miR-17-5p. This miRNA, in a recent study, has been found to be upregulated in plasma of ADPKD patients and exhibited positive correlation with rapid disease progression (Lai *et al*., 2024). The observed upregulation of hsa-miR-20a-5p in our study suggests its involvement in driving abnormal cellular proliferation and cyst expansion, processes that are hallmarks of ADPKD. hsa-miR-27b-3p targets *PIK3CA*, *TFAP2B*, and *EGFR*, which are components of the PI3K-Akt signaling pathway, a crucial mediator of cellular growth and survival. The PI3K-Akt pathway has been implicated in ADPKD pathogenesis through its role in abnormal cell proliferation and apoptosis resistance, both of which are critical for cyst development. The elevated expression of miR-27b-3p, therefore, aligns with previous findings that this pathway is involved in ADPKD, further supporting its contribution to cystogenesis. hsa-miR-3613-3p, in the network analysis was found to target the highest number of genes, including *CDC73*, *CTNNB1*, *WWTR1*, *KIF3A*, *SEC63*, *SET*, *ALG9*, *FLCN*, *IFT20*, and *EP300,* likely having a broad regulatory impact by influencing multiple pathways involved in cell adhesion, proliferation, and signaling simultaneously. *CTNNB1* plays an important role in the Wnt signaling pathway, essential for cell proliferation and differentiation, with abnormalities in this pathway being associated with cystogenesis. *WWTR1* is part of the Hippo signaling pathway, which regulates organ size and tissue growth, while *SEC63* and *KIF3A* are involved in protein trafficking and ciliary function, both of which are disrupted in PKD. *FLCN* (folliculin) is connected to cellular growth and energy sensing, and its mutation has been implicated in renal cyst formation (Chen *et al*., 2015). These pathways are vital for maintaining tissue structure and function, and their dysregulation contributes to cyst formation. The miR-181a-5p, which targets *NOTCH2*, *KIF3A*, and *BCL2*, is associated with the Notch signaling pathway and apoptosis, both critical for maintaining kidney cellular balance. The increased expression of miR-181a-5p hints at its role in promoting cystic growth by affecting cell death and survival mechanisms.

Another interaction from the network was observed with hsa-miR-20a-3p, which targets *TMEM67*, a gene linked to ciliopathies and thus potentially affecting ciliary dysfunction in PKD. Likewise, hsa-let-7i-5p targets *NRAS* and *NPHP3*, genes involved in signal transduction and nephronophthisis respectively, suggesting a role in modulating signaling pathways that contribute to cystogenesis. The hsa-miR-133b specifically targets *BICC1*, a gene crucial for cyst formation and renal development (Tran *et al*., 2010, 2024). The regulation of *BICC1* by miR-133b suggests a significant post-transcriptional control mechanism at play in PKD. Similarly, hsa-miR-205-5p was found to regulate *NOTCH2* and *BICC1*, linking it to cellular differentiation processes and renal tissue organization. In our network analysis, *BICC1* was identified as a target of several miRNAs, and research indicates that *BICC1* works together by binding with *PKD1* and *PKD2* (Tran *et al*., 2024). Bicc1, an RNA-binding protein, plays a crucial role in renal gluconeogenesis, loss of Bicc1 and mutations in *BICC1* is reported to worsen disease severity (Rothé *et al*., 2015, 2020; Tran *et al*., 2024), highlighting its significant role in modifying ADPKD progression.

The pathway enrichment analysis revealed several highly significant pathways affected by miRNA dysregulation in ADPKD that can be crucial for understanding the disease mechanism. The KEGG pathways associated with the identified miRNAs include ubiquitin-mediated proteolysis, the cell cycle, focal adhesion, and autophagy. Dysregulation of the ubiquitin-proteasome system has been previously implicated in ADPKD, where it contributes to the aberrant turnover of polycystin proteins, thereby promoting cystogenesis. The enrichment of the cell cycle pathway, G2M checkpoint and E2F targets suggest that these miRNAs may act as key modulators of uncontrolled epithelial cell proliferation in ADPKD. Dysregulated cell cycle control is a well-established feature of PKD (Lantinga-van Leeuwen *et al*., 2005; Lee, Gusella and He, 2021). Protein processing in endoplasmic reticulum and FoxO signaling pathway suggests that miRNAs may affect protein folding and stress responses, which are essential for cellular integrity in PKD. Pathways such as PKA, AMPK, MAPK, and HIF-1 signaling pathway could reveal how metabolic and hypoxic conditions influence PKD progression. The dysregulation of the cAMP-PKA pathway is a known driver of increased fluid secretion and cyst expansion (Ding *et al*., 2021; Sun *et al*., 2024). The enrichment in cell cycle regulation, apoptotic processes, protein phosphorylation, and autophagy. These processes are tightly regulated in normal renal physiology, but their dysregulation is a hallmark of cystic development. Recent studies have highlighted the multifaceted role of autophagy in ADPKD, suggesting it may contribute to cyst expansion, serve as a compensatory response to cellular stress, or act as a driving factor in the abnormal cellular processes observed in cystic cells (Ponticelli, Moroni and Reggiani, 2023). The enrichment of the ubiquitin-ligase complex and RISC complex suggests miRNA-mediated regulation of protein degradation and post-transcriptional gene silencing. The involvement of the PI3K-Akt and mTOR, and and insulin signaling are critical regulators of cell growth and metabolism, and their hyperactivation in ADPKD has been well documented. Also, mTORC1 has been directly implicated in promoting cyst growth (Margaria *et al*., 2020; Zhang *et al*., 2021; Grahammer *et al*., 2024). Inhibition of the mTOR pathway has shown promise in slowing cyst growth in animal models (Yamaguchi *et al*., 2024). Further, the enrichment of EMT pathway in our analysis indicates that these miRNAs potentially drive fibrosis in ADPKD. The EMT is a process where epithelial cells lose their polarity and acquire mesenchymal characteristics, contributing to fibrosis, a key feature in the progression of ADPKD (Fragiadaki, Macleod and Ong, 2020). Specific miRNAs such as miR-21 and miR-192 have been linked to TGF-beta-induced EMT and fibrosis (Kim *et al*., 2019; Miguel, 2022). These findings suggest that miRNAs targeting all these pathways can serve as potential therapeutic targets in the future.

The integration of miRNA-target gene analysis with PKD-associated genes from GeneCard and OMIM again highlighted several genes critical to renal cyst development. Specifically, the functional enrichment analysis identified 33 genes linked to renal cystogenesis, including *TSC2, PKD1, PKD2, NOTCH2, INVS, NEK8, FLCN,* and *PKHD1,* among others. These genes are known to play key roles in ADPKD pathogenesis, with *PKD1* and *PKD2* mutations being central to cyst formation and disease progression (Cornec-Le Gall et al., 2018). Further, the top 20 hub genes from the miRNA-target gene interaction network, including *NOTCH2, CCND1, TP53, TSC2, PIK3CA*, and *AKT1*, highlight their involvement in cellular processes such as proliferation, apoptosis, and signaling, all of which are disrupted in ADPKD (Harris & Torres, 2018). The identification of these hub genes further suggests that they could serve as key regulatory nodes in the molecular pathways of cyst formation and disease progression and can be further explored.

The observation that multiple miRNAs target common genes within these pathways further strengthens the notion that miRNA-mediated dysregulation plays a crucial role in ADPKD. By targeting multiple components within critical signaling networks, these miRNAs may act as master regulators, amplifying the effects of genetic mutations in *PKD1* and *PKD2*. This provides a mechanistic link between miRNA dysregulation and the abnormal cellular behaviors observed in ADPKD, including increased proliferation, resistance to apoptosis, and abnormal cell polarity.

Our findings suggest that miRNAs regulate diverse cellular functions relevant to the progression of cytogenesis. While some miRNAs might exacerbate disease processes, others may have protective effects, depending on their target genes and the cellular environment. A key question arising from this study is whether the upregulation of miRNAs in blood of ADPKD is a driving factor of disease progression, or a consequence of the cystic environment. Meaning, could these miRNAs actively contribute to cyst formation by targeting crucial genes involved in cellular proliferation, apoptosis, and differentiation, or might some of them have a protective effect, acting to regulate or counterbalance disease progression? Or, is their overexpression a response to the altered cellular environment and stress associated with the disease pathology? Nonetheless, this study emphasizes miRNAs’ potential in ADPKD as both biomarkers and therapeutic targets. Given their involvement in multiple pathways central to cyst formation and disease progression, miRNAs represents a promising area for therapeutic interventions. An innovative approach to restore normal gene expression and signaling could be to target particular miRNAs in order to delay or stop the disease progression.

### Future Direction

The future studies could involve validating the functional roles of the identified key miRNAs in ADPKD by conducting experiments through *in vitro* and *in vivo* models. This might involve testing how increasing or decreasing the levels (e.g., using miRNA mimics or inhibitors) of these miRNAs affects cyst growth and polycystin protein levels. Targeting these miRNAs could offer new treatment options for ADPKD. Also, analysis of the pathways and integration with proteomics and metabolomics data could help better understand how these miRNAs contribute to the cystogenesis and disease progression.

## CONCLUSION

The four miRNAs, hsa-miR20a-5p, hsa-miR27b-3p, hsa-miR3613-3p, and hsa-miR181a-5p, are upregulated in blood sample of the ADPKD patients. The miRNA-gene network, enrichment and expression analysis provides insights into the molecular crosstalk that drives cyst growth and fibrosis, two hallmarks of ADPKD. The upregulation of key miRNAs and their involvement in pathways associated with cell proliferation, apoptosis, autophagy, EMT, and cellular responses to hypoxia suggests that these miRNAs are critical regulators of cystogenesis, consistent with existing literature on the molecular mechanisms of ADPKD. Our findings contribute to a growing body of evidence that miRNAs are important players in the pathogenesis of ADPKD, providing new possibilities for research and potential therapeutic development.

## STATEMENT AND DECLARATIONS

## Data Availability

All data produced in the present study are available upon reasonable request to the authors

## Acknowledgements

We acknowledge the Senior Research Fellowship provided by the Indian Council of Medical Research (ICMR), India, to the first and second authors. We acknowledge Dr. Garima Jain, MPDF, Centre for Genetic Disorders, I.Sc., Banaras Hindu University, India, for her insights and encouragement during the study. We extend our gratitude to Banaras Hindu University, Varanasi, India, for providing the internet essential for conducting this study.

