## Supplementary file 1 for "Role of MicroRNAs in Autosomal Dominant Polycystic Kidney Disease: Expression Profiles and Pathway Analysis"

**Table 1: List of miRNAs linked to PKD and their Source literature**

| **SN** | **miRNA** | **Sample** | **Article’s PubMed ID** |
| --- | --- | --- | --- |
| 1. | miR-17~92  (miR-17  miR-18a  miR-20a) | Mouse model | 23759744 |
| 2. | miR-16 | Patients plasma | 33783812 |
| 3. | miR-107  miR-199a-3p | GEO database |  |
| 4. | mir-143(2)  let-7i(1)  mir-3619(1)  mir-1(4)  mir-133b(2)  mir-205(1)  mir-223(1)  mir-199a(3)  mir-199b(1)  miR-1(2)  miR-133a(2)  hsa-miR-133a(2)  hsa-miR-1(2)  hsa-miR-671  hsa-miR-378  hsa-miR-221  hsa-miR-98  cluster-hsa-mir-223(1)  cluster-hsa-mir-142(1)  cluster-hsa-mir-143(2)  cluster-hsa-mir-133b(2)  cluster-hsa-mir-652(1)  cluster-hsa-mir-338(1)  cluster-hsa-mir-450a-1(4)  cluster-hsa-mir-199a-1(3)  cluster-hsa-mir-199b(1)  cluster-hsa-mir-582(1)  cluster-hsa-mir-3613(1)  cluster-hsa-mir-1-1(4)  cluster-hsa-mir-618(1)  cluster-hsa-mir-2115(1)  cluster-hsa-mir-873(2)  cluster-hsa-mir-2355(1)  cluster-hsa-mir-551a(1)  cluster-hsa-mir-139(1)  cluster-hsa-mir-370(1) | **Primary culture** of ADPKD kidney cells,  **Urine** | 24489795 |
| 5. | miR-17 family  (miR-20a-5p  miR-93-5p  miR-106a-5p)  miR-27a-3p  miR-20a-5p  miR-16-5p  hsa-miR-451a  hsa-miR-16-5p  hsa-miR-223-3p  hsa-miR-21-5p  hsa-miR-20a-5p  hsa-miR-103a-3p  hsa-miR-24-3p  hsa-miR-126-3p  hsa-miR-320a  hsa-miR-320b  hsa-miR-150-5p  hsa-let-7g-5p  hsa-miR-484  hsa-miR-424-5p  hsa-let-7d-3p  hsa-miR-423-5p  hsa-miR-27a-3p  hsa-miR-25-3p  hsa-let-7i-5p  hsa-let-7a-5p  hsa-miR-22-3p  hsa-miR-30e-5p  hsa-miR-29c-3p  hsa-miR-148a-3p  hsa-miR-146a-5p  hsa-miR-30c-5p  hsa-miR-151a-Sp  hsa-miR-423-3p  hsa-miR-26b-5p  hsa-miR-181a-5p  hsa-let-7f-5p  hsa-miR-125a-5p  hsa-miR-199a-5p  hsa-miR-99a-5p  hsa-miR-144-5p  hsa-miR-210  hsa-miR-532-3p | Serum | 34900055 |
| 6. | miR-17~92  miR-21  miR-200  miR-214  miR-185  miR-146b  miR-503  miR-34a  miR-10  miR-204  miR-488 | Book chapter | NBK373371 (BOOK chapter) |
| 7. | miR-21  miR-193  miR-214 |  | 31982550  Review |
| 8. | miR-15A |  | Review |
| 9. | miR-192-5p  miR-194-5p  miR-30a-5p  miR-30d-5p  miR-30e-5p  hsa-mir-1307  hsa-mir-130a  hsa-mir-140  hsa-mir-148  hsa-mir-192  hsa-mir-194-1  hsa-mir-194-2  hsa-mir-197  hsa-mir-25  hsa-mir-27b  hsa-mir-30a  hsa-mir-306  hsa-mir-300  hsa-mir-300  hsa-mir-424  hsa-mir-454  hsa-mir-532  hsa-mir-577  hsa-mir-589  hsa-mir-93  hsa-mir-99a | human urinary exosome miRNA | 32622528 |
| 10. | miR-214 | ADPKD mouse models and cystic kidneys from humans with ADPKD | 32182218 |
| 11. | miR-192  miR-194 | Renal cyst tissue samples from ADPKD tissue.  Mouse  MDCK | 30332302 |
| 12. | miR-31  miR-217  hsa-miR-147  hsa-miR-346  hsa-miR-196a  hsa-miR-126b  hsa-miR-372  hsa-miR-379  hsa-miR-3020-star  hsa-miR-446  hsa-miR-217  hsa-miR-136  hsa-miR-148a  hsa-miR-128a  hsa-miR-377  mmu-miR-380-3p  hsa-miR-3020  hsa-miR-31  hsa-miR-20  hsa-miR-126-star  hsa-miR-30a-3p  hsa-miR-203  hsa-miR-181b  hsa-miR-302b-star  mmu-miR-7b  hsa-miR-99a  hsa-miR-335  hsa-miR-96  hsa-miR-216  hsa-miR-34b  hsa-miR-214  hsa-miR-185 | PKD and control animals | 19102782 |
| 13. | miR-25-3p  miR-92a-3p  miR-3907  miR-18a-5p  miR-27a-3p  miR-128-3p  miR-145-5p  miR-202-3p  miR-224-5p  miR-365a-3p  miR-629-3p  miR-1587  miR-3689-5p  miR-3911  miR4286  miR-4296  miR-4516  miR-4732-5p  miR1260a  miR-1587  miR-21-5p  miR-3907  miR-3911  miR-92a-3b  miR-3907 | Serum,  Venous blood ADPKD patients | 28558802 |
| 14. | miR-17  miR-21  miR-143  miR-223 | Urine | 37060340 |
| 15. | miR-132-3p | ADPKD mouse model, human cell lines. | 33408288 |
| 16. | miR-199a-5p | ADPKD tissues and cell lines | 25588980 |
| 17. | miR-20b-5p  miR-106a-5p | Mice |  |
| 18. | miR-15a | cell line PCK-CCL, which is derived from the PCK rat, | 18949056 |
| 19. | miR501-5p  miR-601  hsa-miR368  hsa-miR601  hsa-miR142-5p  hsa-miR202  hsa-miR196b  miR199a-5p  mir-193b-3p | Fresh cystic kidney tissues were collected from three different ADPKD patients with endstage renal disease (ESRD) | 29323708 |
| 20. | miR-365-1 | Bioinformatic analysis  miRanda | 22411058 |

**Figure: PKD linked 108 common genes in "OMIM" and "GeneCard":**

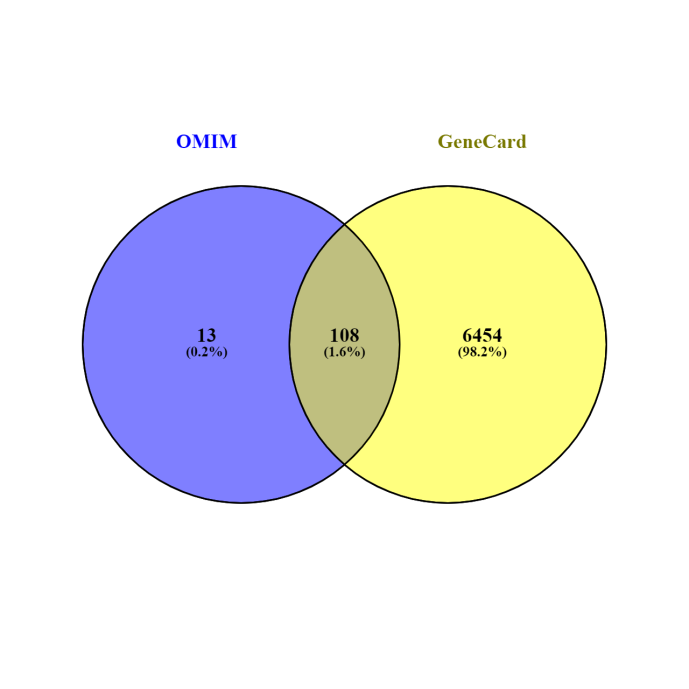

*NRAS, NOTCH2, HAX1, MUC1, CDC73, CYS1, PDIA6, BUB1, DIS3L2, CTNNB1, NPHP3, DZIP1L, WWTR1, PIK3CA, DNAJB11, FGFR3, PTPN13, PKD2, TLR2, NEK1, CASP3, CPLANE1, APC, KIF3A, PKD2L2, KIAA0319, TNF, HSD17B8, MEP1A, TFAP2B, PKHD1, MCM3, SEC63, PKD1L1, EGFR, BRAF, NOS3, TMEM67, RAD54B, PKHD1L1, DNAAF11, GLIS3, ANKS6, INVS, SET, TSC1, OPTN, ITGA8, BICC1, KIF11, PKD2L1, TCF7L2, TRPM5, KCNQ1OT1, CDKN1C, LGR4, GANAB, LRP5, CCND1, ALG8, ALG9, TMEM218, STAT6, IFNG, IFT88, ALG5, MLH3, AKT1, BUB1B, SSTR5, RPL3L, TBL3, GFER, TSC2, PKD1, PGP, E4F1, DNASE1L2, RNPS1, CCNF, NTN3, ATP6V0C, PDPK1, ZNF423, PKD1L3, PKD1L2, DNAAF1, TP53, FLCN, IFT20, NEK8, HNF1B, HOXB-AS1, BCL2, STK11, MCOLN1, PRKCSH, B9D2, BAX, SRC, AURKA, LAMA5, CHEK2, EP300, PKDREJ, PRKX, OFD1, RP2.*

**Table 2: Primer Designing for Stemloop RTqPCR:**

| SN | **miRNA** | **miRBaseAccession no.** | **miRNA ntd Sequence_miRBase** | **Stem Loop RT primer Sequence (5'-->3')** | **SL Length (bp)** | **qPCR Primer** | **PCR/qPCR Sequence (5'-->3')** | **Length (bp)** | **GC (%)** | **Tm (°C)** |
| --- | --- | --- | --- | --- | --- | --- | --- | --- | --- | --- |
|  | **-** | **-** | **-** | **-** | **-** | **RP** | **GTCGTATCCAGTGCAGGGT** | **19** | **57.89** | **59.68** |
| 1 | hsa-miR-3613-3p | MIMAT0017991 | ACAAAAAAAAAAGCCCAACCCUUC | **GTCGTATCCAGTGCAGGGTCCGAGGTATTCGCACTGGATACGACGAAGGG** | 50 | **FP** | **AACACGTGACAAAAAAAAAAGCCC** | **24** | **37.5** | **60.67** |
| 2 | hsa-miR-181a-5p | MIMAT0000256 | AACAUUCAACGCUGUCGGUGAGU | **GTCGTATCCAGTGCAGGGTCCGAGGTATTCGCACTGGATACGACACTCAC** | 50 | **FP** | **AACACGCAACATTCAACGCT** | **20** | **45** | **60.19** |
| 3 | hsa-miR-20a-5p | MIMAT0000075 | UAAAGUGCUUAUAGUGCAGGUAG | **GTCGTATCCAGTGCAGGGTCCGAGGTATTCGCACTGGATACGACCTACCT** | 50 | **FP** | **AGCGAGGCTAAAGTGCTTATAGT** | **23** | **43.48** | **60.21** |
| 4 | hsa-miR-27b-3p | MIMAT0000419 | UUCACAGUGGCUAAGUUCUGC | **GTCGTATCCAGTGCAGGGTCCGAGGTATTCGCACTGGATACGACGCAGAA** | 50 | **FP** | **AACGCACTTCACAGTGGCTA** | **20** | **50** | **60.47** |
| **FP: Forward Primer, RP: Reverse Primer** | | | | | | | | | | |
